# Prevalence and Predictors of Silent Vertebral Compression Fractures: A Cross-Sectional Population-Based Study Using UK Biobank Imaging Data

**DOI:** 10.1101/2025.10.31.25339216

**Authors:** David John Wilson, Richard William Whitehouse, Georgina Marian Allen, Fiona Saunders, Iain Wilson, Richard Abel

**Author notes:** Corresponding Author: Dr David Wilson MBBS BSc FRCP FRCR, St Luke’s Radiology Oxford Ltd, Latimer Road, Oxford OX3 7PF. Contributors DW designed the study, analysed the data, and drafted the manuscript. RW and GA independently reviewed 100 cases for the interobserver reliability study. IW advised on statistical methods and guided R programming. FS retrieved and collated the UK Biobank imaging and health data. RA advised on study design and contributed to manuscript preparation. All the authors read and approved the final version of the manuscript. DW is the guarantor for the study. He accepts full responsibility for the work and the conduct of the study. Declarations of Interest DW and GA are Directors of St Luke’s Radiology Oxford Ltd., a private and NHS outsourced radiology practice registered with the Care Quality Commission. Role of the funding source The lead investigator (DW) self-funded this study using personal resources from his clinical practice. The authors have no financial or commercial interest in the results of this study. Ethics approval UK Biobank has full ethical approval from the National Information Governance Board for Health and Social Care and the North-West Multi-Centre Research Ethics Committee (11/NW/0382). Permission to access and analyse UK Biobank data for this study was approved under UKB application number 17295. The Imperial College London Bone Lab holds regular Patient in Research meetings to discuss the nature and findings of the project with the patient panel. This project was discussed at a meeting in 2019. Data Sharing This study used data from the UK Biobank AUGMENT study, available to approved researchers through the standard UK Biobank access process (application number 17295). Data were obtained from the UK Biobank under application 17295. Further access is subject to approval by the UK Biobank. Declaration All authors have read and approved the revised manuscript David Wilson FRCP - 30 October 2025 Richard Whitehouse MD - 30 October 2025 Georgina Allen FRCP - 30 October 2025 Fiona Saunders PhD - 30 October 2025 Iain Wilson PhD - 30 October 2025 Richard Abel PhD - 30 October 2025.

## Abstract

**Objectives:** To estimate the prevalence of silent vertebral compression fractures (VCF) in an asymptomatic population and to assess the demographic and clinical predictors using data from the UK Biobank.

**Design:** A cross-sectional study using artificial intelligence-assisted six-point morphometry of dual-energy x-ray absorptiometry spine images.

**Setting:** UK Biobank imaging study at Stockport clinical research facility.

**Participants:** 2 446 asymptomatic volunteers aged 40 to 79 years with no history of spinal trauma or vertebral fracture.

**Main outcome measures:** VCFs were defined as greater than or equal to20 percent height loss, measured by artificial intelligence, with manual correction of annotations and confirmation by a musculoskeletal radiologist. Association with age, sex, bone mineral density, body mass index, and back pain were analysed.

**Results:** Of 2 446 participants, 763 (31.1 percent) had at least one vertebra with > 20 percent height loss. Fracture prevalence increased with age: 26.8 percent (37 of 138) in those aged 40-49 years, 26.1 percent (196 of 752) at 50-59 years, 33.0 percent (380 of 1 153) at 60-69 years, and 37.2 percent (150 of 403) at 70-79 years. Excluding mild deformities, 18 percent of those aged 40-49 years had moderate or severe fractures (> 25 percent height loss). Fractures were 11.5 percent more common in men than in women. Age was a significant predictor, but bone mineral density (BMD) and body-mass index (BMI) were not. Predictive models showed limited performance (sensitivity 44.4 percent, specificity 68.2 percent). Interobserver agreement was substantial (Fleiss kappa = 0.74).

**Conclusions:** Silent VCFs are frequent by midlife, including among younger adults with normal bone density. Men are affected more often than women, and conventional risk factors inadequately identify those at risk. Earlier detection through opportunistic or population imaging may enable timely intervention and inform future screening and fracture-prevention strategies.

**Key messages:**

a. What is already known on this topic VCFs are common but often unrecognised, particularly when asymptomatic. Most evidence is derived from older or symptomatic populations.
b. What this study adds Silent VCFs affect more than 30 percent of adults and over one-quarter of those under 50. They are more frequent in men and often occur despite normal bone mineral density. Conventional risk factors show limited predictive value.
c. How this study might affect research, practice or policy Expanding fracture assessment to younger populations could enable earlier intervention and inform future screening guidelines.

Manuscript

## Introduction

Vertebral compression fractures (VCF) are common, with around 66,000 symptomatic cases identified annually in England and Wales, many incidentally detected during investigations for unrelated conditions. (1;2) Many patients are unaware that they have sustained a VCF and recall no acute back spine pain or trauma. (3) Asymptomatic fractures are often longstanding and may herald further fractures with height loss, functional decline, and increased mortality. Early medical management may improve outcomes.

Studies using Dual Energy X-ray Absorptiometry (DXA), radiographs, CT, or MRI have produced widely varying estimates of VCF prevalence. The European Vertebral Osteoporosis Study (EVOS) found a prevalence of 12-20 percent, depending on the morphometric criteria used. (4-6) Other papers reported prevalence rates ranging from 4 percent to more than 60 percent, with higher rates in Scandinavian and hospital-based studies (7-12). Although fractures are traditionally considered a post-menopausal condition, several studies have identified comparable or higher rates in men (8-10), challenging conventional sex-specific assumptions. Prospective work from the Study of Osteoporotic Fractures (SOF) clarified definitions of incident vertebral deformity and highlighted ascertainment challenges, but focused mainly on clinically apparent events rather than truly silent prevalent deformities (13). A meta-analysis of asymptomatic women showed that many with vertebral fractures had non-osteoporotic bone density (43 percent osteopenic; 32 percent normal BMD), demonstrating the limited predictive value of BMD alone (14).

Most previous research has focused on symptomatic or elderly populations, leaving uncertainty about the actual burden and predictors of silent VCFs in middle-aged adults. This heterogeneity impedes the development of evidence-based pathways for opportunistic or population screening. Standardised reviews of asymptomatic populations are needed to guide early intervention.

This study addresses that gap by analysing UK Biobank participants who underwent whole-spine DXA imaging. We aimed to quantify the prevalence of silent VCFs in asymptomatic adults aged 40-79 years, and identify demographic and clinical predictors, including age, sex, body mass index (BMI), bone mineral density (BMD), and back pain, using artificial-intelligence-assisted morphometry verified by an expert radiologist. By applying uniform imaging criteria in a large, population-based sample, we sought to provide a reproducible estimate of the hidden vertebral fracture burden and to determine whether clinical evidence can reliably identify individuals at risk.

Understanding who sustains these fractures and when they occur has implications that extend beyond clinical radiology. Early detection of silent VCFs could enable timely initiation of bone-protective therapy, inform fracture-prevention guidelines, and shape future public-health screening policies aimed at reducing the substantial morbidity and healthcare costs associated with spinal fragility fractures.

## Methods

### Study cohort

The UK Biobank is a prospective population study with half a million participants, 100,000 of whom were invited for whole-spine DXA imaging with vertebral fracture analysis as part of an imaging clinic assessment. This study, ethically approved under application 17295, also includes health assessments and medical records under a data access agreement.

DXA scans (GE iDXA, software v16.00.158) were performed at the Stockport Clinical Research Facility (2014-2016) from T4-L4, excluding poor-quality levels. Available data included demographics, DXA parameters, medical history, and prescriptions; individuals with prior spinal trauma or vertebral fractures were excluded. Height and weight were measured using standard procedures, while other variables were collected via a touchscreen questionnaire. Lumbar T-scores were automatically calculated by the iDXA software, and BMI was derived from height and weight.

We analysed 2 466 volunteers who had no history of spine fracture or trauma. Power calculations identified whether comparisons were adequately powered, limited, or underpowered based on the minimum detectable difference (MDD) at 80 percent power, as shown in Table 1.

**Table 1.** Fracture prevalence and power assessment by demographic and clinical characteristics.

| Variable | Category | Fractures (n/N, %) | MDD at 80% power (pp) | Power Assessment |
| --- | --- | --- | --- | --- |
| **Age Group** | **40-49** | **37/138 (26.8%)** | **Reference** |  |
| Age Group | 50-59 | 196/752 (26.1%) | 11.5 | Limited |
| Age Group | 60-69 | 380/1153 (33.0%) | 11.2 | Limited |
| Age Group | 70-79 | 150/403 (37.2%) | 12.2 | Limited |
| **Sex** | **male** | **429/1151 (37.3%)** | **Reference** |  |
| Sex | female | 334/1295 (25.8%) | 5.5 | Adequate |
| **Ethnicity** | **British** | **705/2238 (31.5%)** | **Reference** |  |
| Ethnicity | African | 2/6 (33.3%) | 53.2 | Underpowered |
| Ethnicity | Other | 52/178 (29.2%) | 10.1 | Limited |
| Ethnicity | South Asian | 4/24 (16.7%) | 26.7 | Underpowered |
| **BMI Group** | **Normal** | **244/895 (27.3%)** | **Reference** |  |
| BMI Group | Underweight | 2/10 (20.0%) | 39.7 | Underpowered |
| BMI Group | Overweight | 336/1053 (31.9%) | 5.7 | Adequate |
| BMI Group | Obese | 181/488 (37.1%) | 7.0 | Adequate |
| **BMD** | **Normal** | **532/1724 (30.9%)** | **Reference** |  |
| BMD | Osteopaenia | 167/553 (30.2%) | 6.3 | Adequate |
| BMD | Osteoporosis | 37/102 (36.3%) | 13.2 | Limited |
| **Simple analgesics** | **FALSE** | **501/1574 (31.8%)** | **Reference** |  |
| Simple analgesics | TRUE | 262/872 (30.0%) | 5.5 | Adequate |
| **NSAID** | **FALSE** | **537/1686 (31.9%)** | **Reference** |  |
| NSAID | TRUE | 226/760 (29.7%) | 5.7 | Adequate |
| **Opiate** | **FALSE** | **746/2405 (31.0%)** | **Reference** |  |
| Opiate | TRUE | 17/41 (41.5%) | 20.4 | Underpowered |
| **Calcium** | **FALSE** | **742/2394 (31.0%)** | **Reference** |  |
| Calcium | TRUE | 21/52 (40.4%) | 18.2 | Underpowered |
| **Anti resorptive** | **FALSE** | **739/2383 (31.0%)** | **Reference** |  |
| Anti resorptive | TRUE | 24/63 (38.1%) | 16.5 | Underpowered |
| **Vitamin D** | **FALSE** | **672/2157 (31.1%)** | **Reference** |  |
| Vitamin D | TRUE | 91/289 (31.5%) | 8.1 | Adequate |
| **Steroid** | **FALSE** | **757/2428 (31.1%)** | **Reference** |  |
| Steroid | TRUE | 6/18 (33.3%) | 30.7 | Underpowered |
| **Back pain in last 3 months** | **TRUE** | **172/565 (30.4%)** | **Reference** |  |
| Back pain in last 3 months | FALSE | 591/1881 (31.4%) | 6.2 | Adequate |

**Table 2.** Unadjusted and adjusted odds ratios for vertebral fracture by demographic and clinical characteristics.

| Variable | Category | Unadjusted OR<br>(95 % CI) | P (level, unadj) | Adjusted OR<br>(95 % CI) | P (level, adj) | P (global) |
| --- | --- | --- | --- | --- | --- | --- |
| **Age_Group** | **40-49** | **1.00 (Ref)** |  | **1.00 (Ref)** |  | **0.000** |
| Age_Group | 50-59 | 0.96 (0.64-1.47) | 0.854 | 0.96 (0.63-1.49) | 0.847 |  |
| Age_Group | 60-69 | 1.34 (0.91-2.02) | 0.146 | 1.27 (0.85-1.95) | 0.252 |  |
| Age_Group | 70-79 | 1.62 (1.06-2.50) | 0.027 | 1.44 (0.93-2.27) | 0.112 |  |
| **Sex** | **male** | **1.00 (Ref)** |  | **1.00 (Ref)** |  | **0.000** |
| Sex | female | 0.58 (0.49-0.69) | 0.000 | 0.59 (0.49-0.71) | 0.000 |  |
| **Ethnicity** | **British** | **1.00 (Ref)** |  | **1.00 (Ref)** |  | **0.378** |
| Ethnicity | African | 1.09 (0.15-5.58) | 0.923 | 0.90 (0.12-4.74) | 0.909 |  |
| Ethnicity | Other | 0.90 (0.64-1.25) | 0.527 | 0.93 (0.65-1.30) | 0.661 |  |
| Ethnicity | South Asian | 0.43 (0.13-1.15) | 0.130 | 0.42 (0.12-1.13) | 0.116 |  |
| **BMI_Group** | **Normal** | **1.00 (Ref)** |  | **1.00 (Ref)** |  | **0.002** |
| BMI_Group | Underweight | 0.67 (0.10-2.68) | 0.610 | 0.54 (0.08-2.29) | 0.454 |  |
| BMI_Group | Overweight | 1.25 (1.03-1.52) | 0.026 | 1.19 (0.97-1.47) | 0.094 |  |
| BMI_Group | Obese | 1.57 (1.24-1.99) | 0.000 | 1.65 (1.28-2.12) | 0.000 |  |
| **BMD** | **Normal** | **1. (Ref)** |  | **1.00 (Ref)** |  | **0.480** |
| BMD | Osteopaenia | 0.97 (0.79-1.19) | 0.770 | 1.07 (0.85-1.33) | 0.575 |  |
| BMD | Osteoporosis | 1.28 (0.83-1.92) | 0.252 | 1.60 (1.02-2.48) | 0.037 |  |
| **Simple analgesics** | **FALSE** | **1.00 (Ref)** |  | **1.00 (Ref)** |  | **0.361** |
| Simple analgesics | TRUE | 0.92 (0.77-1.10) | 0.362 | 0.90 (0.74-1.09) | 0.279 |  |
| **NSAID** | **FALSE** | **1.00 (Ref)** |  | **1.00 (Ref)** |  | **0.295** |
| NSAID | TRUE | 0.91 (0.75-1.09) | 0.296 | 0.91 (0.75-1.12) | 0.385 |  |
| **Opiate** | **FALSE** | **1.00 (Ref)** |  | **1.00 (Ref)** |  | **0.162** |
| Opiate | TRUE | 1.58 (0.83-2.93) | 0.156 | 1.61 (0.83-3.06) | 0.151 |  |
| **Calcium** | **FALSE** | **1.00 (Ref)** |  | **1.00 (Ref)** |  | **0.157** |
| Calcium | TRUE | 1.51 (0.85-2.63) | 0.151 | 1.54 (0.80-2.90) | 0.185 |  |
| **Anti resorptive** | **FALSE** | **1.00 (Ref)** |  | **1.00 (Ref)** |  | **0.239** |
| Anti resorptive | TRUE | 1.37 (0.81-2.28) | 0.233 | 1.24 (0.66-2.24) | 0.494 |  |
| **Vitamin D** | **FALSE** | **1.00 (Ref)** |  | **1.00 (Ref)** |  | **0.909** |
| Vitamin D | TRUE | 1.02 (0.78-1.32) | 0.909 | 1.12 (0.84-1.49) | 0.419 |  |
| **Steroid** | **FALSE** | **1.00 (Ref)** |  | **1.00 (Ref)** |  | **0.845** |
| Steroid | TRUE | 1.10 (0.38-2.85) | 0.844 | 1.06 (0.36-2.79) | 0.912 |  |
| **Back pain in last 3 months** | **TRUE** | **1.00 (Ref)** |  | **1.00 (Ref)** |  | **0.660** |
| Back pain in last 3 months | FALSE | 1.05 (0.85-1.29) | 0.660 | 1.06 (0.85-1.32) | 0.603 |  |

We examined associations between vertebral fractures and several participant-level variables: age group (40-49, 50-59, 60-69, 70-79 years), sex (male, female), back pain (yes, no), lumbar-spine T-score (normal, osteopenia, osteoporosis), body-mass-index (BMI) category (underweight, normal, overweight, obese), vitamin D supplementation (yes, no), calcium supplementation (yes, no), and anti-resorptive therapy (yes, no). For each variable, fracture prevalence was expressed as the proportion of participants with at least one fracture within each category. Reference groups were defined a priori (for example, males for sex, normal BMI for body mass index).

Fractures were defined as a greater than or equal to 20 percent reduction in vertebral height and classified as mild (20-25 percent), moderate (25-40 percent), or severe (>40 percent). (15) Unlike semiquantitative grading, the exact percentage compression for each vertebra was calculated to minimise bias and over-classification. Fracture detection used a validated six-point morphometric algorithm (IoMSpine, Optasia Medical, Manchester, UK) that measured vertebral body heights and computed wedge, biconcave, and crush ratios. Automated annotations were manually reviewed to exclude errors due to vertebral tilt, scoliosis, Schmorl’s nodes, poor image quality, or congenital anomalies. Each case was assessed by a radiographer and independently verified by an experienced musculoskeletal radiologist (DW, 45 years’ experience). This combined automated and expert workflow ensured standardised and reproducible assessment of vertebral fractures in an asymptomatic cohort.

### Interobserver study

Agreement was assessed in 100 randomly selected cases (1,287 vertebrae) using Cohen’s and Fleiss’ weighted Kappa statistics, interpreted per Landis and Koch.(16)

### Statistical methods

Statistical analyses were performed in R (version 4.3.0), using RStudio (2023.06.0+421). Normality was tested (Anderson-Darling, Shapiro-Wilk, Kolmogorov-Smirnov); as most variables were non-normal, non-parametric and categorical tests were used (Table S1). Accordingly, χ^2^ tests were used for categorical variables, and Spearman’s, Pearson’s, Wilcoxon, and Kendall tests for continuous variables, as appropriate. Logistic regression was applied to binary outcomes, and Poisson regression for count data.

Associations between continuous variables such as BMD and the presence of vertebral fractures were assessed using multivariable logistic regression to estimate adjusted odds ratios, as this approach allows study of independent effects while controlling for potential confounders.

We trained random-forest models (ntree = 500, mtry = √p) with 10-fold cross-validation. Predictor variables included age, sex, BMI, BMD, back pain, and medication use; the binary outcome was presence or absence of greater than or equal to20 percent vertebral height loss. (17) Logistic regression models were fitted with all predictors. Model performance was assessed by ROC AUC analysis, sensitivity and specificity.

Interobserver agreement was tested using Cohen’s weighted Kappa and Fleiss’ Kappa. Visualisations were produced using ggplot2. Additional methodological and validation details are provided in the Supplementary Appendix.

## Results

### Cohort Demographics

Of 2 466 participants, 20 were excluded due to prior spine injury, leaving 2 446 for analysis. Of 32 058 vertebrae, 220 (0.69 percent) were excluded due to poor image quality.

### Prevalence of Silent Fractures

Among 2 446 participants, 763 (31.1 percent) had fractures greater than or equal to20 percent compression, affecting 26.0 percent of women and 37.3 percent of men (Table 1). In non-fractured vertebrae, mean mid-height increased from T4 to L3 by 0.70-0.89 mm (3.5-5.5 percent) per level, then decreased by 0.80 mm (−3.4 percent) at L4. Sex, back pain in the three months before the DXA examination, and high BMI were all significantly associated with fractures (p < 0.0001); however, age, BMD, and medication were not.

### Fractures in Younger Populations and Age Association

Vertebral fracture prevalence increased with age. In the 40-49 year group, 26.8 percent (37 of 138) had fractures and served as the reference. Prevalence was similar in the 50-59 year group at 26.1 percent (196 of 752; odds ratio 0.96, 95 percent confidence interval 0.64 to 1.47; p = 0.85) and higher in the 60-69 year group at 33.0 percent (380 of 1153; odds ratio 1.34, 95 percent confidence interval 0.91 to 2.02; p = 0.15), though neither difference was statistically significant. Age showed a graded increase in fracture prevalence, and only those aged 70-79 years 37.2 percent (150 of 403) had significantly higher odds (odds ratio 1.62, 95 percent confidence interval 1.06 to 2.50; p = 0.03) (Figures 1-3).

**Figure 1.**
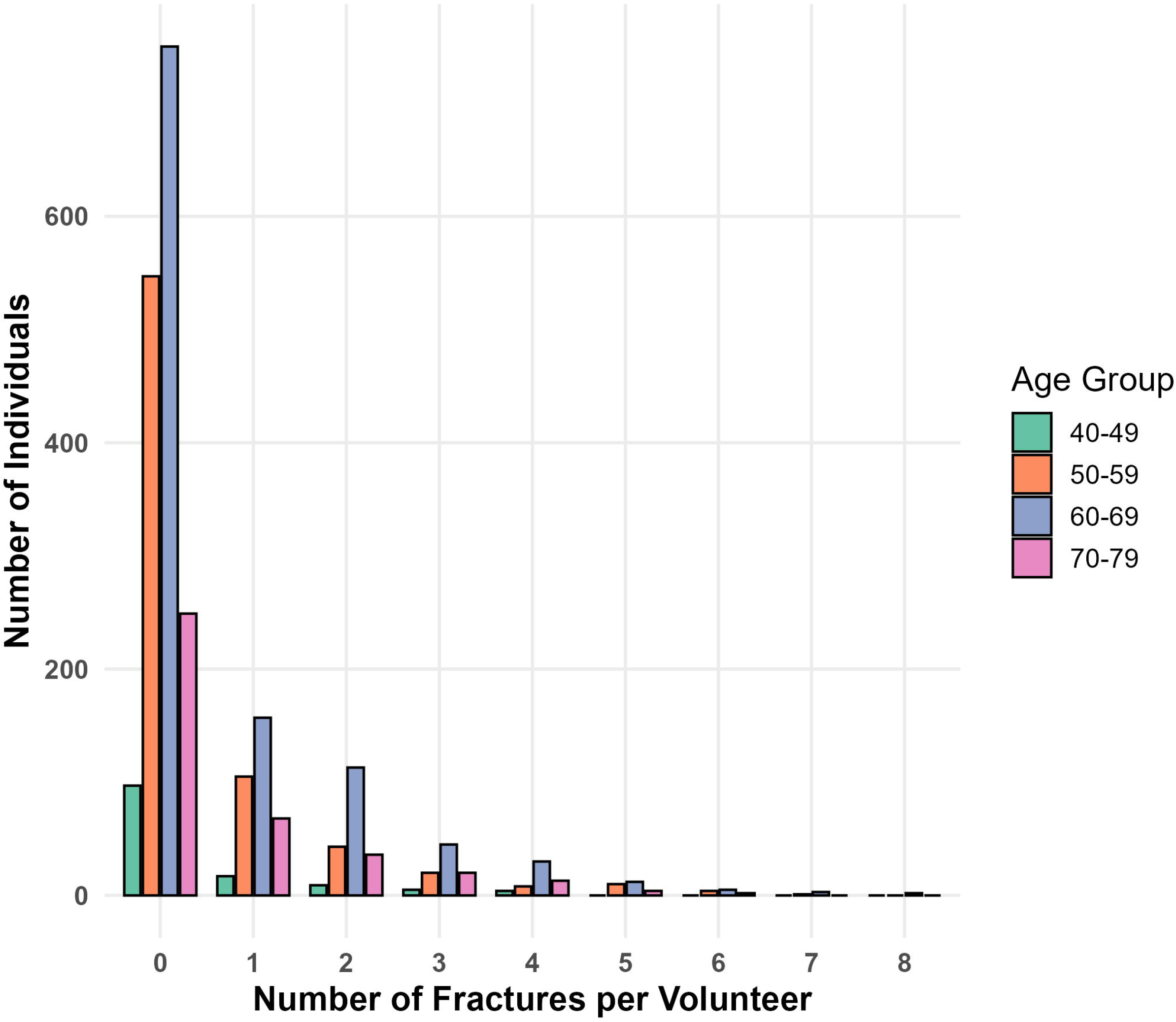
Distribution of vertebral fracture counts by age group. Each bar represents the number of individuals with 0 to 8 vertebral fractures in each decade cohort from age 40 to 79 years. Fracture burden increased with age, and a higher proportion of older participants had multiple fractures.

Moderate fractures were frequently observed in the 40-49 group, while severe fractures occurred more often in middle-aged (50-59) than older individuals (ρ = −0.11, p < 0.0001; Figure 3) (Figure S2, Supplementary Appendix).

### Sex differences

Fractures were more prevalent in men across all age groups, and the gap narrowed with age (Figure 3, Table 1).

### Multiple Fractures

Multiple fractures per individual increased with age; only 4.9 percent of those under 60 had greater than or equal to2 fractures, compared with >10 percent in the 70-79 group (Figures 1 and 2).

**Figure 2.**
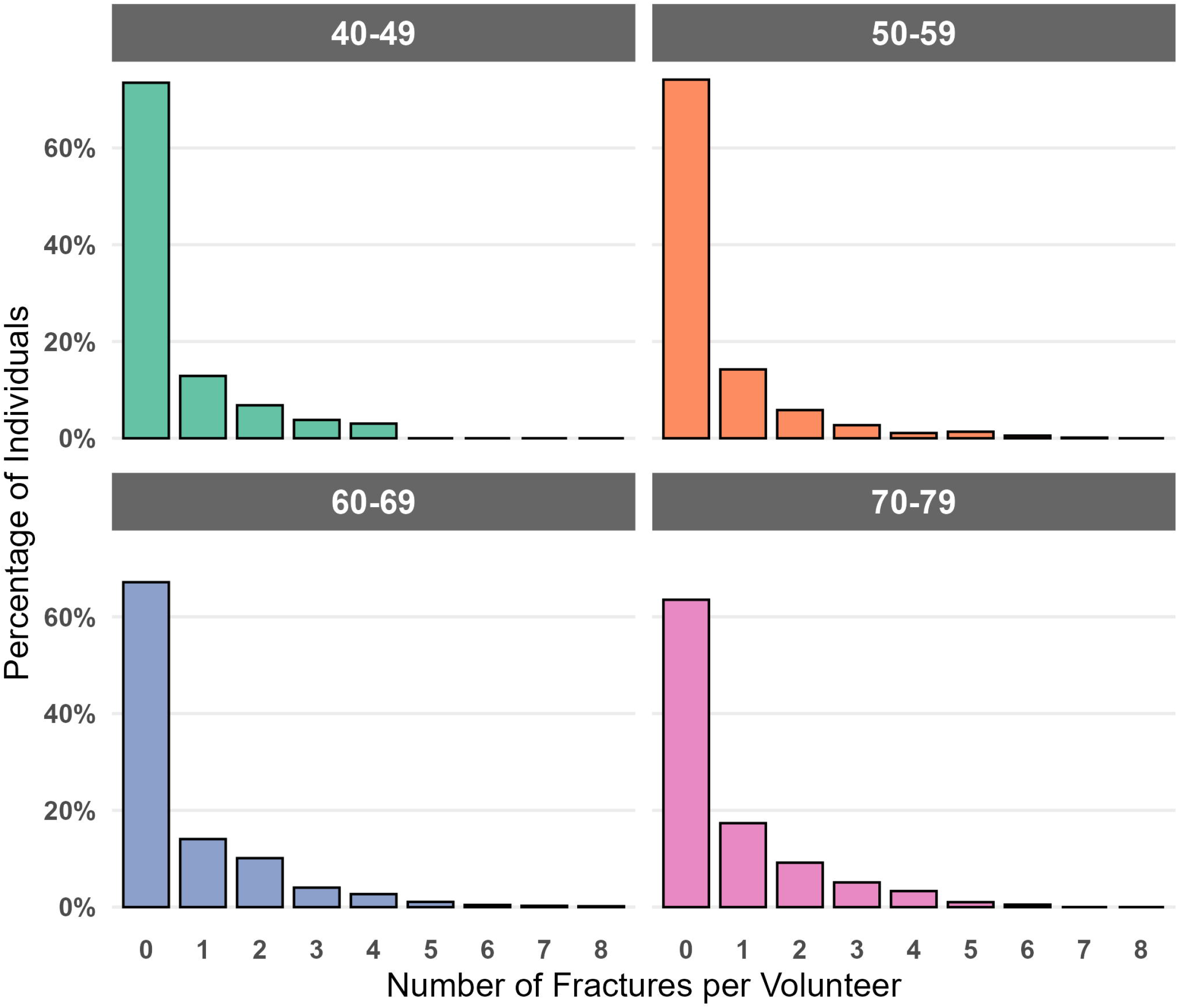
Proportion of individuals in each age group by number of vertebral fractures. Bar heights represent the percentage of individuals within each age group who have a given number of fractures. Distributions are normalised within groups to illustrate how fracture burden is spread across ages.

**Figure 3.**
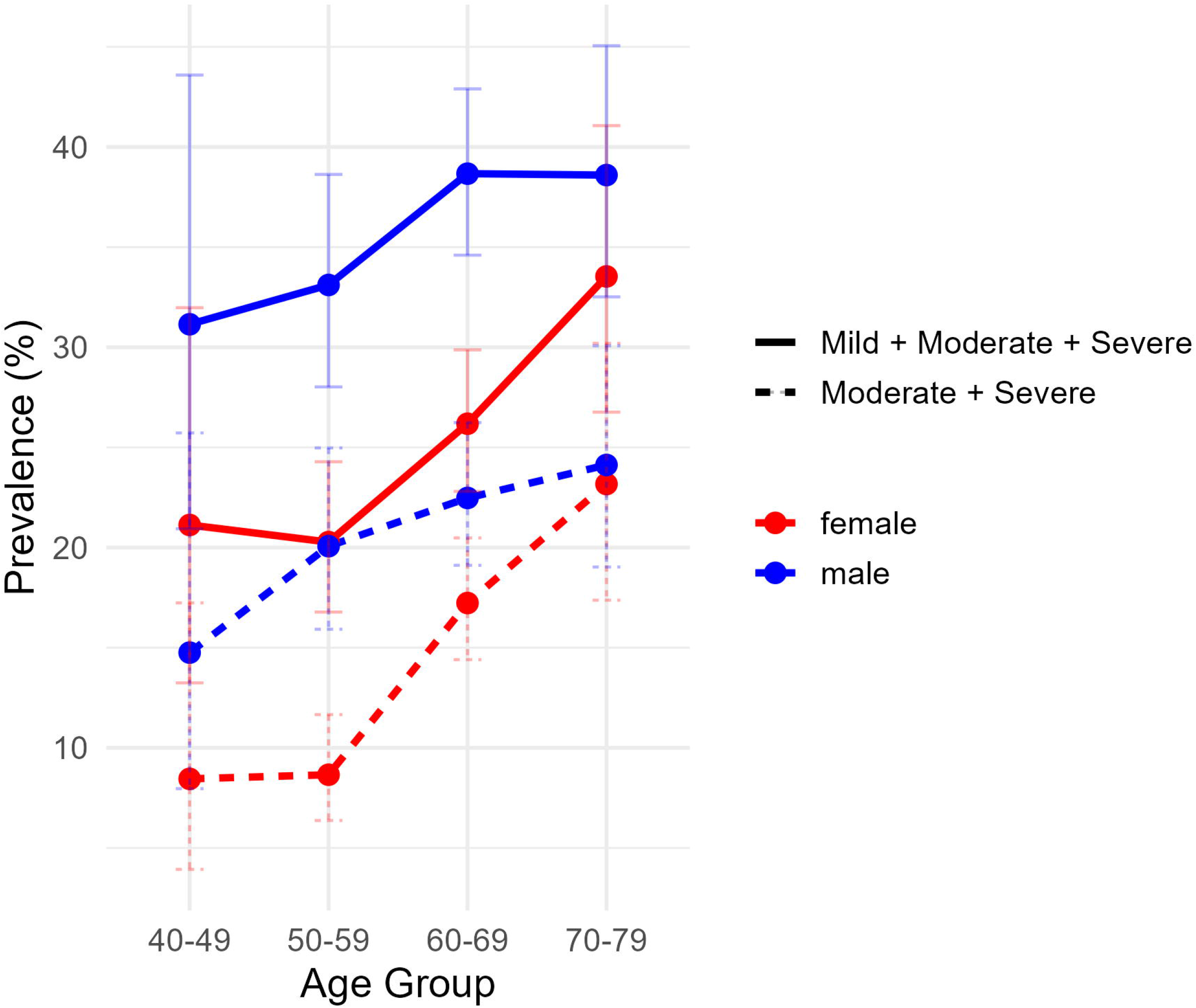
Prevalence of fractures in each age group divided by sex. The upper graph includes all fractures, the lower (dotted) graph excludes the mild fractures. Fractures increase in frequency with age, and in these asymptomatic volunteers, show slightly higher prevalence in men for all but the oldest age group.

### Back pain association

Recent back pain (within 3 months) was reported by 23.1 percent. Odds of fracture were significantly higher in this group OR 2.42; 95 percent CI: 1.86-3.15, p < 0.001 (Table 1).

### Bone mineral density and BMI associations

Median BMD values differed slightly between participants with and without vertebral fractures, but the distributions largely overlapped, indicating similar value ranges between the groups. In multivariable logistic regression, BMD was not significantly associated with vertebral fracture risk (adjusted OR = 1.13, 95 percent CI 0.92-1.39, p = 0.250).

Participants with fractures had significantly higher median compression than those without Wilcoxon rank-sum W = 589 813, p < 0.0001. Because the compression values were non-normally distributed (Table S1), a nonparametric test was used for the unadjusted comparison, while multivariable logistic regression was applied separately to estimate adjusted odds ratios for fracture risk.

### Prediction Modelling

Random forest and logistic regression models showed poor predictive power (sensitivity: 44.4 percent, specificity: 68.2 percent; χ^2^ = 0.658, p = 0.417). No meaningful improvements were observed with cross-validation or age stratification.

### Artificial intelligence misclassification and alternative diagnoses

Across all 31 798 vertebrae assessed, the artificial intelligence initially identified 4 590 (14.4 percent) vertebrae as fractured; 606 (13.2 percent) were subsequently reclassified after manual review as benign alternative findings such as spondylosis, diffuse idiopathic skeletal hyperostosis (DISH), or Schmorl’s nodes, representing 1.9 percent of all vertebrae examined. The remaining 3,984 artificial intelligence false positives (12.5 percent of all vertebrae) showed no alternative diagnosis, likely reflecting vertebral shape variation, spinal tilt or algorithmic threshold sensitivity.

Degenerative processes (spondylosis, DISH, osteophyte formation) accounted for approximately two-thirds of all alternative diagnoses, while developmental variants (Schmorl’s nodes, block vertebrae, cupid’s bow) comprised around one-quarter. False-positive classifications were most frequent in the mid-thoracic region (T8-T9), corresponding to normal physiological wedging and areas of frequent degenerative change. Annotator misclassifications were rare (<2 percent) and confined mainly to the lumbar spine.

### Radiologist Interobserver reliability

The weighted Fleiss Kappa was 0.74, indicating substantial agreement. Observer-specific patterns were similar, with minimal discrepancies (maximum 2.6 percent at T11) (Table S2, Figure S1 Supplementary Appendix).

## Discussion

### Study Overview

This study is, to our knowledge, the first to use a fully standardised, reproducible method combining automated six-point morphometry, manual verification, and expert radiologist review to identify VCFs in asymptomatic individuals. Because no universal in vivo gold standard exists, we defined fractures asgreater than or equal to20 percent reduction in vertebral height, consistent with the Genant semiquantitative approach and other large population studies. Automated morphometric analysis, exclusion of anatomical variants, and independent expert review maximised diagnostic specificity, providing the most robust available framework for large-scale epidemiological assessment.

Using whole-spine DXA imaging in 2 446 asymptomatic UK Biobank participants, we found a high prevalence of silent VCFs (30 percent), with higher rates in men, frequent occurrence in younger adults, and poor predictability from conventional risk factors. Unlike earlier studies based on retrospective or clinically indicated imaging, our design reduces selection bias and provides a standardised population reference for vertebral fracture burden.

### Prevalence and Distribution of Silent Fractures

The 30 percent prevalence we observed exceeds most prior estimates, EVOS reported 12-20 percent, CaMoS 21-23 percent, and CT-based studies 4-20 percent.(4-6;8;9;11;12) This likely reflects our more sensitive, standardised approach combining AI-assisted six-point morphometry with expert review. Broader T4-L4 coverage and inclusion of asymptomatic volunteers may also reveal earlier or subclinical disease.

These methodological advances suggest that vertebral fractures are more common than previously recognised and highlight the need for population-based screening strategies.

### Fractures in Younger Populations

Over one-quarter of participants under 50 had vertebral fractures, some moderate or severe. These findings challenge the view that such fractures are confined to older age and suggest that many deformities seen later in life originate decades earlier. Earlier detection could allow intervention before symptomatic disease develops.

### Sex Differences

Contrary to traditional assumptions linking s predominantly to postmenopausal women, we found consistently higher fracture rates in men across all age groups (37.0 percent in men versus 25.6 percent in women). This 11 percent greater prevalence in men challenges existing paradigms and suggests that silent fractures may exhibit different demographic patterns compared to symptomatic ones. Previous studies, such as those by Felsenberg et al., 2002, (18) showed equal fracture prevalence in younger cohorts, with later divergence; however, our findings suggest a male predominance even in asymptomatic populations. This pattern warrants further investigation into potential genetic, anatomical, or lifestyle factors that might explain this disparity.

### Multiple Fractures

A notable proportion of participants had multiple fractures, with 4.9 percent of those under 60 having two or more. The presence of multiple fractures, even in younger individuals, suggests systemic disorders affecting vertebral integrity rather than isolated traumatic injury, such as multiple metastases, myeloma, or renal osteodystrophy. (19)In asymptomatic individuals, it raises the possibility of cumulative microtrauma during events or developmental factors compromising vertebral structure without triggering clinical symptoms.

### Association with Back Pain

Although participants reported no history of spinal trauma, 23 percent had back pain within the preceding three months. Fractures were significantly more frequent in this group, indicating that many “silent” deformities may cause mild or transient symptoms that remain unrecognised. It is challenging to distinguish asymptomatic from minimally symptomatic fractures in population studies.

### Distribution of Fracture Severity

Fracture severity showed a weak negative correlation with age: moderate deformities were commonest in the 40-49 group, while severe fractures predominated among older adults. Severe changes in younger people may reflect unrecognised high-energy injury, whereas remodelling over time could obscure severe compression in older individuals.

### Relationship with Bone Mineral Density and BMI

Although fracture prevalence correlated weakly with BMD and BMI (p < 0.001), many affected individuals had normal bone density, confirming that BMD alone does not adequately capture risk. This supports evidence that bone architecture and quality influence strength as much as density. The positive association between BMI and fracture prevalence may relate to altered biomechanics or adiposity-related inflammation rather than traditional osteoporotic mechanisms..(20)

### Artificial intelligence misclassification and vertebral morphology

Although fracture prevalence correlated weakly with BMD and BMI (p < 0.001), many affected individuals had normal bone density, confirming that BMD alone does not adequately capture risk. This aligns with evidence that bone architecture and quality influence strength as much as density. The positive association between BMI and fracture prevalence may relate to altered biomechanics or adiposity-related inflammation rather than traditional osteoporotic mechanisms.

### Prediction of Fractures

Despite testing multiple predictors, including age, BMI, BMD, back pain, and medication use, both random-forest and logistic models showed limited discrimination (sensitivity 44 percent, specificity 68 percent). Conventional risk factors appear inadequate for identifying individuals with silent fractures, revealing an important gap in fracture prediction and prevention.

### Ethnicity and socioeconomic status

Despite testing multiple predictors, including age, BMI, BMD, back pain, and medication use, both random-forest and logistic models showed limited discrimination (sensitivity 44 percent, specificity 68 percent). Conventional risk factors appear inadequate for identifying individuals with silent fractures, revealing an important gap in fracture prediction and prevention.

## Study Limitations

The UK Biobank cohort comprises motivated volunteers and may not reflect the full UK population. The cross-sectional design prevents the determination of fracture timing or progression. Limited data on back-pain severity and duration constrain the interpretation of symptom-fracture relationships. Although our combined automated and expert workflow enhances specificity, the absence of a histological gold standard means that some misclassification of vertebral shape changes is unavoidable.

## Conclusion

Silent VCFs are common, affecting nearly one-third of asymptomatic adults, particularly men and even those under 50. The presence of multiple and moderate-to-severe deformities in younger individuals suggests an earlier onset than previously recognised. Many occur despite normal bone mineral density, raising concerns regarding current risk-screening models.

These findings have major public-health implications. Broader application of vertebral-fracture assessment, including opportunistic assessment during imaging, could enable timely bone-protective therapy and reduce future symptomatic fractures. Longitudinal studies are warranted to define the natural history of these silent fractures and to determine whether early detection mitigates morbidity and the healthcare burden.

## Data Availability

This study used data from the UK Biobank AUGMENT study under approved application [17295]. Data are available to bona fide researchers through the standard UK Biobank access procedures (https://www.ukbiobank.ac.uk/enable-your-research/apply-for-access
).

## Acknowledgements

The late Dr Paul Bromiley (University of Manchester) for his assistance with the artificial intelligence algorithm. Wellcome Collaborative Award (209 233/Z/17/Z) supported Dr Fiona Saunders’ post. Dr John Pinney (Imperial College London) and Dr Paul Bassett provided statistical advice. This paper forms part of a thesis written by DW for the Doctor of Medicine by Research degree at Imperial College London. DW is dyslexic and used Grammarly (Grammarly Inc., San Francisco, CA, USA) to correct spelling and grammar while preparing the manuscript. The text was not prepared using generative artificial intelligence. No professional editorial assistance was used.

## Research in Context

### Evidence before this study

We searched PubMed and Google Scholar for studies published between January 1990 and December 2024. We used combinations of the terms “vertebral compression fracture”, “asymptomatic”, “silent”, “prevalence”, “morphometry”, “population-based”, and “UK Biobank”. Most studies reporting vertebral fracture prevalence were either limited to symptomatic individuals or groups of older adults (>60 years). Asymptomatic volunteer studies reported vertebral fracture prevalence ranging from 12 percent to 23 percent. Most studies used radiographic or semiquantitative methods rather than morphometric vertebral analysis. Few included younger, asymptomatic populations. To our knowledge, none have combined artificial intelligence with expert-verified morphometry in a large, asymptomatic group.

### The added value of this study

This is the first study to assess the prevalence and severity of silent VCFs in a large, asymptomatic population using morphometry on DXA examinations validated by a combination of artificial intelligence and expert review. We found that 31 percent of participants had VCFs, including 25.8 percent of those aged 40-49. We show that severe fractures are not limited to older age groups. Moderate fractures were present in one in five individuals in their 40s. Fractures were more common in men. Predictive modelling with machine learning methods demonstrated poor discrimination. Neither BMD nor high BMI was a reliable predictor of fracture.

### Implications of all the available evidence

Silent VCFs may be identified at a younger age than previously suggested. They are often present in asymptomatic volunteers with normal bone mineral density (BMD). These findings suggest the need for earlier surveillance and screening beyond current BMD-based thresholds.

Fracture prevalence and power assessment by demographic and clinical characteristics. Values are shown as the number of participants with a fracture divided by the total within each category, with percentages in parentheses. MDD = minimum detectable difference in fracture prevalence at 80 percent power. Power assessment classifies each variable as Strong (< 5 percentage points), Adequate (5-10 pp), Limited (10-15 pp), or Underpowered (> 15 pp).

Unadjusted and adjusted odds ratios for vertebral fracture by demographic and clinical characteristics. Values are odds ratios (ORs) with 95 percent confidence intervals (CI) derived from logistic regression. Unadjusted ORs are from single-factor models; adjusted ORs include all variables shown. P (level) represents the Wald test for each level versus its reference; P (global) represents the likelihood-ratio test for the variable overall.

## Supplementary Appendix

Additional data are provided in the Supplementary Appendix, including interobserver reliability analyses (Tables S1–S2) and supplementary figures (Figures S1–S3)

