## Supplementary Appendix for "Prevalence and Predictors of Silent Vertebral Compression Fractures: A Cross-Sectional Population-Based Study Using UK Biobank Imaging Data"

### Supplementary Tables

**Table S1.** Tests for normality of continuous variables using Anderson–Darling statistic
All variables showed non-normal distribution ($p < 0.0001$).

| Variable | A-statistic | p-value | Conclusion |
|---|---|---|---|
| BMI | 20.963 | < 0.0001 | Not normal |
| Age | 15.962 | < 0.0001 | Not normal |
| Weight | 10.629 | < 0.0001 | Not normal |
| Height | 5.1534 | < 0.0001 | Not normal |
| Fractures per volunteer | 394.37 | < 0.0001 | Not normal (extreme deviation) |
| Spine BMD | 1.9176 | < 0.0001 | Not normal |
| Lumbar spine T score | 2.7088 | < 0.0001 | Not normal |
| Max vertebral compression | 62.568 | < 0.0001 | Not normal (severe deviation) |

**Table S2 Kappa statistics for the three pairs of observers.** This shows substantial agreement between the radiologists.

| **Rater pairing** | **Weight Kappa (95% CI)** |
|---|---|
| Pair 1 (RW v DW) | 0.69 (0.62-0.75) |
| Pair 2 (RW v GA) | 0.74 (0.68-0.80) |
| Pair 3 (DW v GA) | 0.70 (0.63-0.76) |

### Supplementary Figures

**Figure S1.** Interobserver agreement in vertebral fracture classification
Percentage disagreement between observers at each vertebral level. The most disputed findings were at T11.

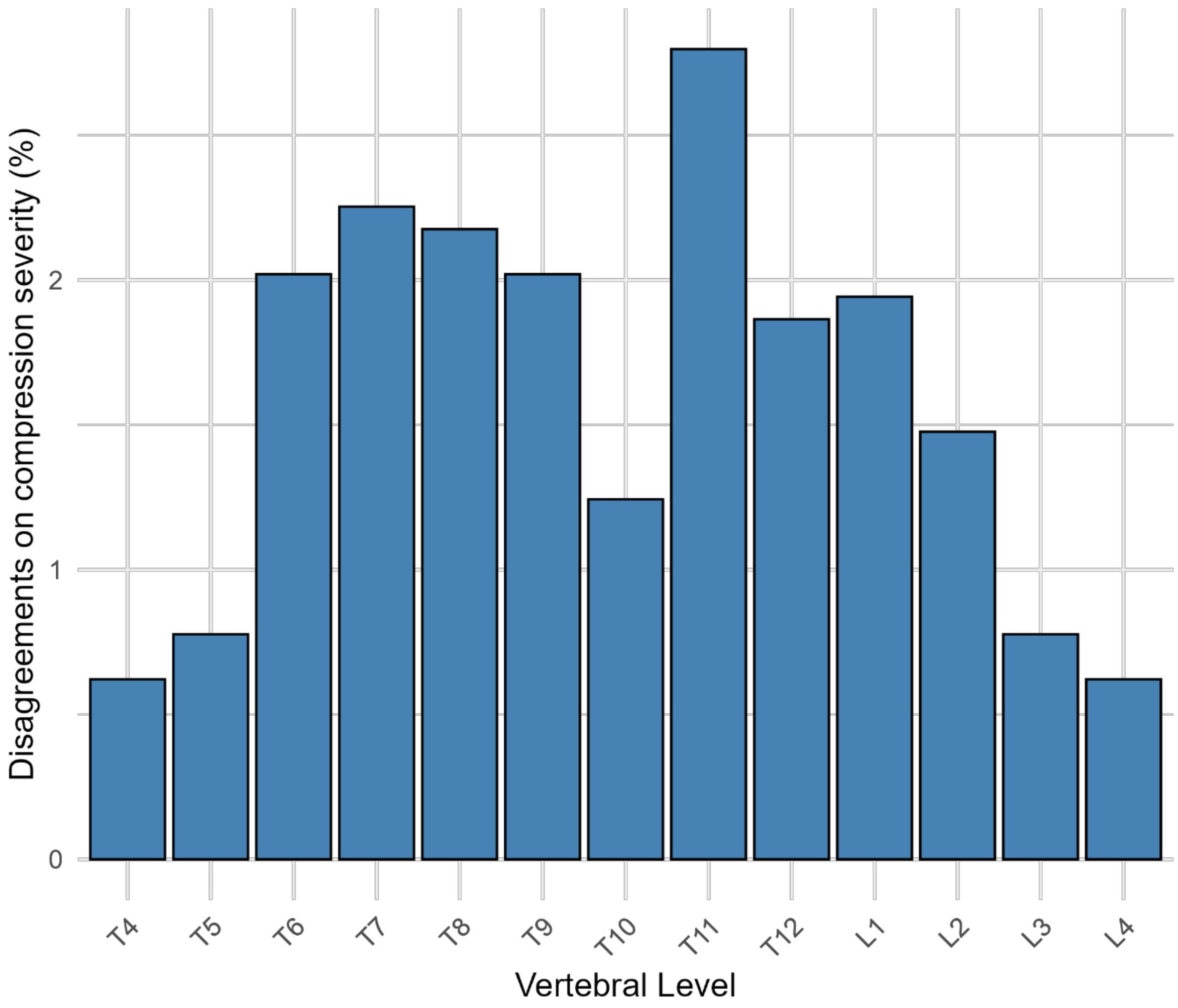

Disagreements on compression severity (%)
0
1
2
T4
T5
T6
T7
T8
T9
T10
T11
T12
L1
L2
L3
L4
Vertebral Level

**Figure S2: Selecting only fractures in the moderate and severe categories (over 25% compression).** The fracture prevalence increased with age. However, the total for the 40-49 group was 18.1%, with the 70-79 group being 29.5%. This suggests that over half the fractures seen in the oldest group may have been present for over 20 years.

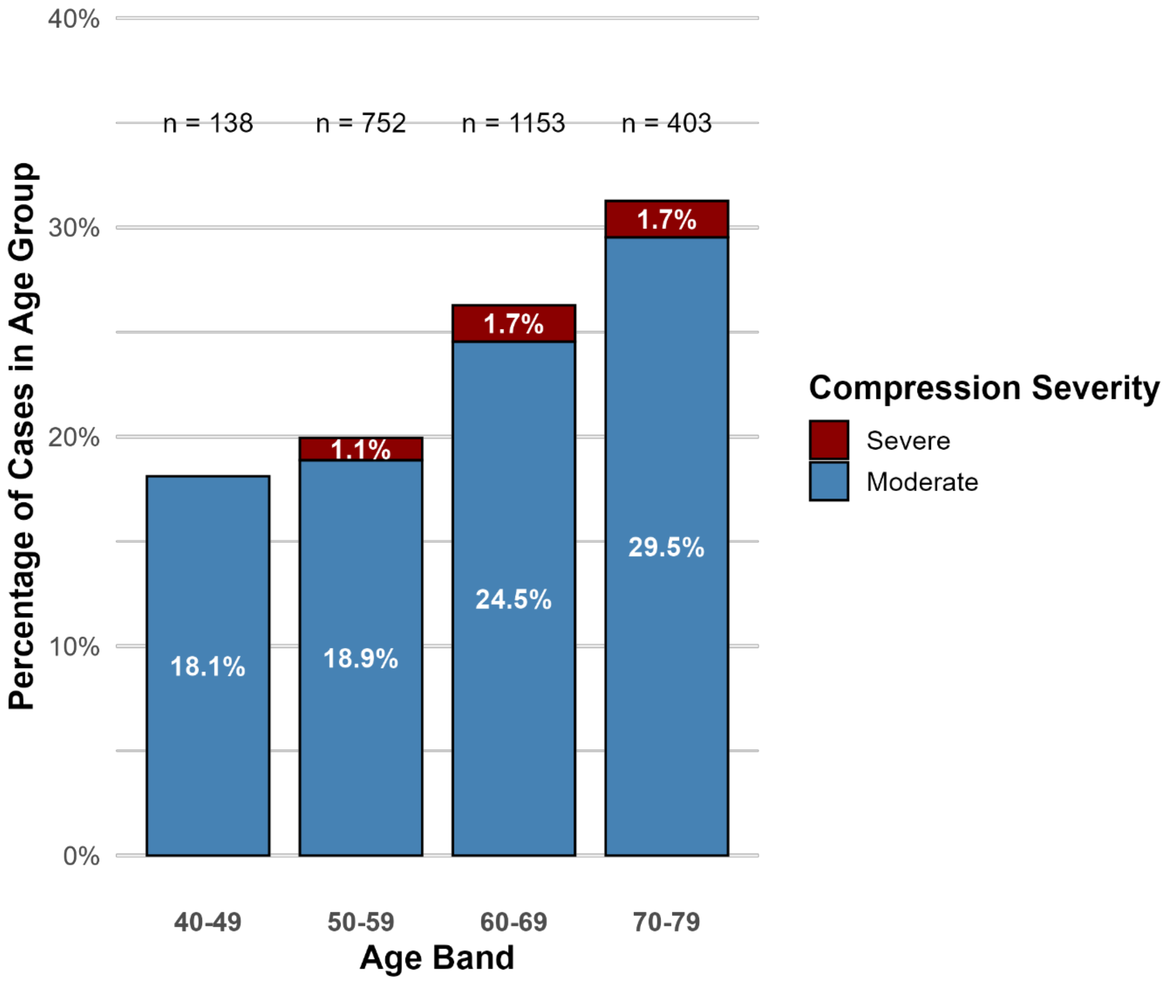

**Figure S3. Distribution of AI false-positive vertebral fracture classifications by vertebral level and alternative diagnosis**

Stacked bar chart showing the number of vertebrae initially labelled as fractured by the artificial-intelligence (AI) system but subsequently judged normal after expert review, grouped by vertebral level (T4–L4) and type of alternative diagnosis. Degenerative findings (spondylosis, diffuse idiopathic skeletal hyperostosis [DISH], osteophyte formation) accounted for Bars represent vertebral counts at each level; colours indicate diagnosis categories.

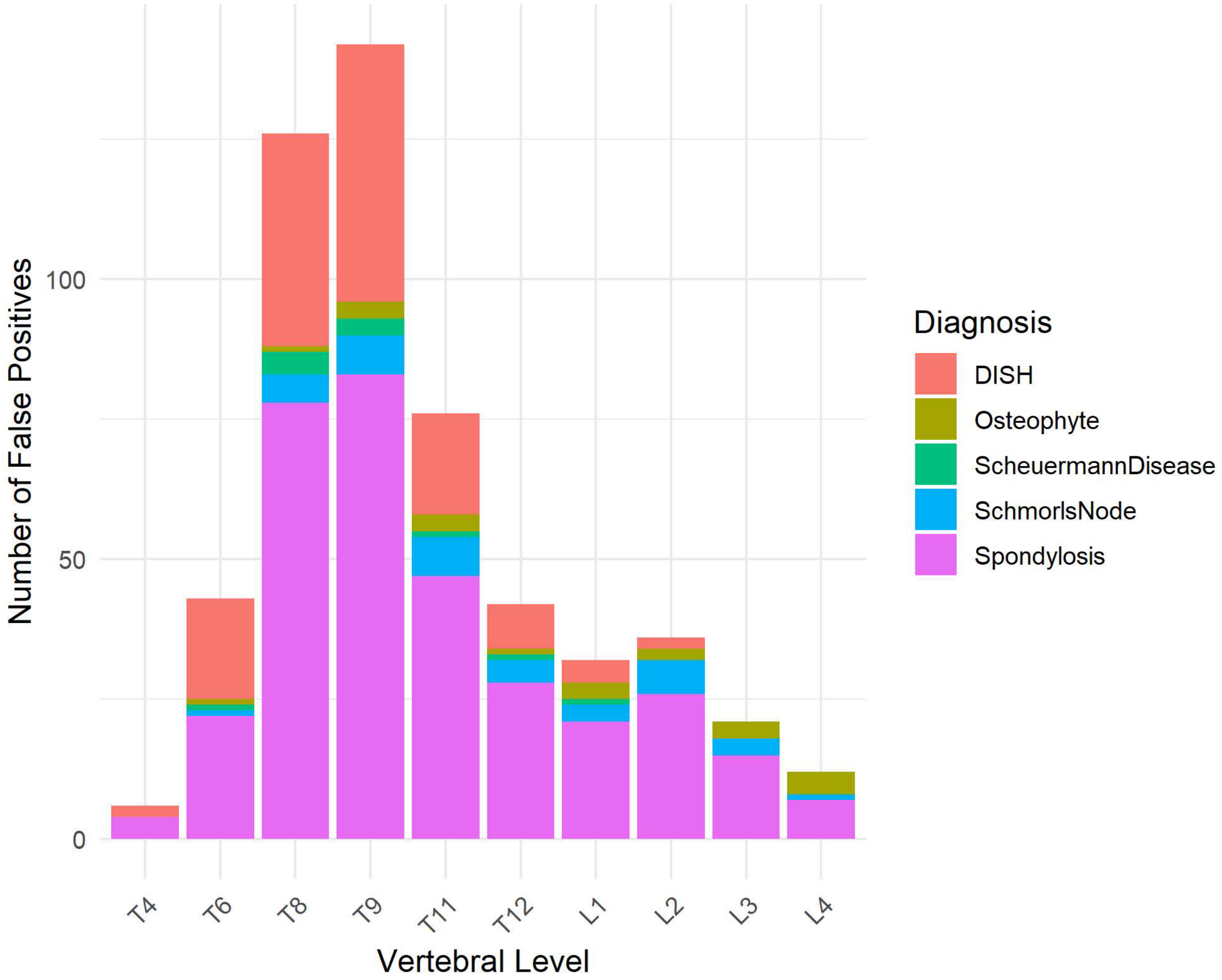
